# Precision MRI phenotyping of muscle volume and quality at a population scale

**DOI:** 10.1101/2023.03.02.23286689

**Authors:** Marjola Thanaj, Nicolas Basty, Brandon Whitcher, Elena P. Sorokin, Yi Liu, Ramprakash Srinivasan, Madeleine Cule, E. Louise Thomas, Jimmy D. Bell

## Abstract

**Introduction:** Magnetic resonance imaging (MRI) enables direct measurements of muscle volume and quality, allowing for an in-depth understanding of their associations with anthropometric traits, and health conditions. However, it is unclear which muscle volume measurements: total muscle volume, regional measurements, measurements of muscle quality: intermuscular adipose tissue (IMAT) or proton density fat fraction (PDFF), are most informative and associate with relevant health conditions such as dynapenia and frailty.

**Methods:** We developed a pipeline to automatically segment and extract image-derived phenotypes (IDPs) including total and regional muscle volumes and measures of muscle quality, and applied it to the neck-to-knee Dixon images in 44,520 UK Biobank participants. We further segmented paraspinal muscle from 2D quantitative MRI to quantify muscle PDFF and iron concentration. We defined dynapenia based on grip strength below sex-specific cut-off points and frailty based on five criteria. We used logistic regression to investigate the association between muscle volume and quality measurements and dynapenia and frailty.

**Results:** Muscle volumes were significantly higher in male compared with female participants, even after correcting for height while, IMAT, (corrected for muscle volume) and paraspinal muscle PDFF were significantly higher in female compared with male participants. From the overall cohort, 7.6% (N = 3,261) were identified with dynapenia, and 1.1% (N = 455) with frailty. Dynapenia and frailty were positively associated with age and negatively associated with physical activity levels. In dynapenia, muscle volume IDPs were most informative, particularly total muscle exhibiting odds ratios (OR) of 0.392 and 95% confidence intervals (CI) = 0.361 - 0.426, while for frailty, muscle quality was found to be most informative, in particular thigh IMAT volume indexed to height squared (OR = 1.396, 95% CI = 1.374 - 1.418), both with p-values below the Bonferroni-corrected threshold (*p* < *8*.*6* × *10*^−*5*^).

**Conclusions:** Our fully automated method enables the quantification of muscle volumes and quality suitable for large population-based studies. For dynapenia, muscle volumes particularly those including greater body coverage such as total muscle are the most informative, whilst, for frailty, markers of muscle quality were the most informative IDPs. These results suggest that different measurements may have varying diagnostic values for different health conditions.

## 1 Introduction

Direct population-scale measurements of muscle have been relatively limited, most studies have used measures of fat-free mass (FFM) by bioelectrical impedance analysis (BIA), or measures of lean body mass (LBM) from dual-energy X-ray absorptiometry (DXA) (Hofsteenge, Chinapaw and Weijs, 2015; Zillikens *et al*., 2017). Unlike direct muscle measures, FFM contains all nonfat components of the body, while LBM additionally contains significant and disparate contributions from organs, skin, bones, body water, and essential fat (Scafoglieri and Clarys, 2018). On the other hand, magnetic resonance imaging (MRI) enables the direct quantification of both muscle mass and quality, with the latter, generally related to levels of muscle fat infiltration (Gallagher et al., 2005; Linge et al., 2018).

Until recently, measurement of total body muscle volume and distribution by MRI has been typically limited to relatively small cohorts, due to the cost and time-consuming requirements of image acquisition and analysis. The lack of automated techniques led researchers to rely on the measurement of single or multiple cross-sectional areas as an index of overall muscle mass, with a single slice at the third lumbar vertebra (L3) (Schweitzer et al., 2015), individual muscles groups such as the iliopsoas (Fitzpatrick et al., 2020) or anatomical groupings such as the ‘thigh muscles’ (Linge *et al*., 2018) being among the most popular approaches. The extent to which proxies of muscle volume can provide sufficient information to discern the overall impact of muscle volume and quality on health is yet to be fully ascertained. Additionally, there is a lack of consensus regarding whether muscle measurements should be used independently, as ratios or whether these should be indexed to height (Linge, Heymsfield and Dahlqvist Leinhard, 2020).

Ageing and morbidity-related declines in physical function have shown to be associated with a loss of muscle mass, strength and reduction in muscle quality (Ungar and Marchionni, 2017; Sizoo et al., 2021). While muscle-related conditions such as sarcopenia and frailty are typically associated with ageing, it is increasingly apparent that multiple long-term factors increase the risk of these conditions at a relatively younger age (Hanlon et al., 2018; Dodds *et* al., 2020). Despite their common association with ageing and multiple diseases, previous studies have shown that even low muscle strength, termed as dynapenia (measured by hand grip strength) is associated with hallmarks of ageing (Jones *et al*., 2021) and is predictive of future long-term morbidity and mortality (Rantanen et al., 1999; Mitchell et al., 2012).

Recent MRI studies have shown the combination of low muscle mass and high levels of muscle fat infiltration to be a predictor of all-cause mortality, suggesting that muscle fat infiltration could contribute to the definition of sarcopenia (Linge *et al*., 2021). However, much of this work has been limited to specific anatomical regions such as the thigh muscle. Although, there is a growing interest in measuring iliopsoas muscle size, primarily because of its potential as a marker of sarcopenia, which indirectly influences conditions associated with sarcopenia and frailty, including adverse health outcomes such as morbidity and mortality (Ebbeling et al., 2014; Kasahara et al., 2017). Furthermore, previous studies have shown that atrophy and changes in fat in paraspinal muscles, stemming from sarcopenia, are linked to functional impairments and chronic back pain (Masaki et al., 2016; Kim et al., 2019), making these muscles important subjects of study in the context of ageing and health.

While the UK Biobank imaging protocol does not encompass the entire body, including only the neck-to-knee region, it is still possible to derive meaningful overall muscle measures including overall muscle volume within that region, specific muscle groups including iliopsoas muscle, thigh muscles, paraspinal muscles, as well as intermuscular adipose tissue (IMAT). In this study, we report both MRI muscle mass and quality measurements for 44,520 UK Biobank participants. We define our muscle volume image-derived phenotypes (IDPs) as total muscle volume, iliopsoas muscle volume, thigh and mid-thigh muscle volume. Additionally, our muscle quality IDPs include thigh IMAT volume, mid-thigh IMAT volume, fat in the paraspinal muscles and the ratio of the mid-thigh IMAT to muscle volume. We further assess which IDPs related to muscle volume and quality, both in their unadjusted form and after adjusting for the most common allometric scaling (height squared), are most informative and associate them with anthropometric factors and relevant health conditions including dynapenia and frailty.

## 2 Materials and Methods

### 2.1 Data

A total of 44,520 UK Biobank participants were included in this analysis. Participant data from the cohort was obtained through UK Biobank Access Application number 44584. The UK Biobank has approval from the North West Multi-Centre Research Ethics Committee (REC reference: 11/NW/0382). All measurements were obtained under these ethics, adhering to relevant guidelines and regulations, with written informed consent obtained from all participants. Researchers may apply to use the UK Biobank data resources and the results generated in this study, by submitting a health-related research proposal that is in the public interest. More information may be found on the UK Biobank researchers and resource catalogue pages (https://www.ukbiobank.ac.uk).

### 2.2 Image processing and MRI measurements

Full details of the UK Biobank magnetic resonance imaging (MRI) abdominal protocol have previously been reported (Littlejohns et al., 2020). The data presented in this paper primarily consisted of two MRI acquisition methods: the chemical-shift-based water-fat separation MRI, commonly known as Dixon, which covered the region from the neck to the knee, and a distinct single-slice multiecho MRI acquisition specific to the liver. All data were analysed using our dedicated image processing pipelines for the Dixon and two types of single-slice multiecho acquisitions: (i) Iterative Decomposition of Water and Fat with Echo Asymmetry and Least-Squares Estimation (IDEAL) and (ii) gradient echo (GRE), with deep learning algorithms employed to segment organs and tissue (Liu *et al*., 2021). Proton density fat fraction (PDFF) and R2* were calculated from the Phase Regularized Estimation using Smoothing and Constrained Optimization (PRESCO) method from both the single-slice IDEAL and GRE acquisitions (Liu et al., 2021).

Our pipeline generates more than 30 image-derived phenotypes (IDPs) from the Dixon MRI data, the subset of IDPs included in this study are primarily muscle volumes, including *total muscle* (within neck-to-knee volume), *thigh (left+right) muscle volume and iliopsoas (left+right) muscle volume*. For training the deep learning model, we utilised 108 manual annotations of total muscle and 151 annotations of the iliopsoas muscles. The Dice similarity coefficients on 20% out-of-sample testing data were 0.94 for total muscle and 0.95 for the iliopsoas muscle segmentations. The thigh muscle volumes were derived from the total muscle segmentations using anatomical landmarks from the femur. Together with these IDPs, to avoid any bias of height we also analysed a 10cm image slab, placed at the midpoint of the length of the femur on each leg. This is referred to as the mid-thigh region throughout the text.

In addition to muscle volume IDPs several adipose tissue depots were extracted, including abdominal subcutaneous adipose tissue (ASAT), visceral adipose tissue (VAT) and intermuscular adipose tissue (IMAT, the tissue located between muscle groups) volumes (Liu et al., 2021). We also obtained a measure of intramyocellular fat stored in the paraspinal muscles, referred to as PDFF to avoid confusion. We finally extracted a measure of paraspinal muscle iron concentration by transforming the R2* to iron concentration in mg/g (Wood et al., 2005). For model training, we had 195 manual annotations of the IDEAL paraspinal muscles and 187 manual annotations of the GRE paraspinal muscles. The Dice similarity coefficient on 20% out-of-sample testing data was 0.81 and 0.86 for the paraspinal muscle segmentations in IDEAL and GRE acquisitions, respectively.

To account for the potential confounding effect of height on muscle and fat volumes, IDPs were indexed to the commonly used allometric scaling, height squared (Linge, Heymsfield and Dahlqvist Leinhard, 2020). Muscle volume was defined for all muscle IDPs including total muscle volume, iliopsoas muscle volume, thigh and mid-thigh muscle volume, whereas muscle quality was defined from all fat measurements including thigh IMAT volume, mid-thigh IMAT volume and paraspinal muscle PDFF. We further evaluate muscle quality measurement related to IMAT, correcting for muscle volume in the mid-thigh. This correction is represented as the mid-thigh IMAT/muscle ratio calculated as follows: (mid-thigh IMAT volume / (mid-thigh IMAT volume + mid-thigh muscle volume)) * 100.

### 2.3 Quality Control

IDPs with insufficient anatomical coverage were reported as missing values and a visual inspection of the MRI data was performed to determine potential extreme values in the IDPs to confirm exclusion. To ensure the full anatomical coverage of the organs a threshold was defined as follows:

- *Total muscle volume*: 5 L < total muscle < 60 L. No participants were identified above or below this threshold.
- *Thigh muscle volume*: 1 L < thigh muscle < 15 L. A total of 25 participants were identified and removed from all subsequent analyses.
- *Thigh IMAT volume*: thigh IMAT > 50 ml. One participant was identified and removed from all subsequent analyses.
- Images from all excluded participants were subsequently visually inspected to confirm the original finding.
- A total of 87 participants were excluded from the volumetric analysis of the Dixon MRI data due to missing values. We excluded 1,547 participants for the muscle and fat volume index analyses, mainly due to missing standing height data. From the single-slice multiecho data 319 participants were excluded from the PDFF and iron concentration analysis in the paraspinal muscle due to missing values.

### 2.4 Phenotype definitions

Anthropometric measurements including age, body mass index (BMI), waist and hip circumferences and hand grip strength (HGS) were taken at the UK Biobank imaging visit. Ethnicity was self-reported at the initial assessment visit and was categorised as follows: White (any British, Irish or other white background); Asian (any Indian, Pakistani, Bangladeshi or any other South Asian background); Black (any Caribbean, African or any other Black background); Chinese and Others (mixed ethnic background including White and Black Caribbean, White and Black African, White and Asian, and other ethnic groups). Sex was self-reported and included those recorded by the NHS and those obtained at the initial assessment visit. The Townsend deprivation index was calculated immediately prior to participants joining the UK Biobank.

Participant’s excess metabolic equivalents of task (MET) in hours per week were computed by multiplying the time in minutes spent in each activity, including walking, moderate and vigorous activity, on a typical day by the number of reported days doing the exercise and the respective MET scores using the methods previously described (O’Donnell *et* al., 2020). Excess METs scores were at 2.3, 3 and 7 for walking, moderate and vigorous activity, respectively (O’Donnell et al., 2020). The total physical activity MET was computed by taking the sum of walking, moderate and vigorous MET in hours per week. Any activity lasting less than 10 minutes on a typical day was recorded to 0 for any of the three categories of activity (walking, moderate and vigorous activity). Additionally, for each of the three categories of activity, values exceeding 1260 minutes per week (equivalent to an average of 3 hours per day) were truncated at 1260 minutes (Bradbury et al., 2017). In our following analysis total MET will be referred to as MET. All physical activity measures, alcohol intake frequency and smoking status were self-reported at the UK Biobank imaging visit.

Relevant conditions of interest including dynapenia and frailty were defined according to published criteria:

- *Dynapenia* was defined as low muscle strength with HGS below 27 kg in male participants and below 16 kg in female participants (Dodds *et al*., 2014, 2020; Silva et al., 2022; Wilkinson et al., 2022). The HGS used was that of the dominant hand. If participants reported using both hands, the mean of the right and left hand was utilised. Initially, we planned a stricter definition of sarcopenia including low muscle quality as recommended by EWGSOP2 (Cruz-Jentoft et al., 2019) which combines both HGS and DXA-measured appendicular lean mass (ALM)/height2: 6.0/7.0 kg/m^2^ (females/males). However, due to the limited availability of UKBB DXA data, (only 4,683 participants had available DXA measurements) resulting in only 0.3% (78M / 22F) of the cohort meeting the full sarcopenia criteria. Hence, our analysis was focused only on dynapenia.
- *Frailty* was defined using the criteria adopted by (Hanlon et al., 2018), to be used with self-reported UK Biobank questionnaire responses which required the presence of 3 out of 5 indicators. These indicators included weight loss, exhaustion (more than half the days or nearly every day), no or only light (once a week or less) physical activity in the last 4 weeks, slow walking speed, and low HGS (Supplementary Table S1). All the frailty indicators were taken at the UK Biobank imaging visit. Additional classifications included pre-frailty (1 or 2 of the 5 indicators), and not frail (none of the indicators).

### 2.5 Statistical Analysis

All summary statistics, hypothesis tests, regression models and figures were performed in R software environment version 4.1.3. Figures were produced using the *ggplot2* package (Wilkinson, 2011). All descriptive characteristics are presented as means with standard deviations (SD) for quantitative variables and as percentages for categorical variables. Spearman’s rank correlation coefficient (ρ) was used to assess monotonic trends between variables. The Wilcoxon rank-sum test was used to compare the means between two groups. One-way ANOVA was used to compare multiple groups.

We explored the association between muscle and fat IDPs and dynapenia by performing a logistic regression analysis model adjusting for age, sex, ethnicity, alcohol intake frequency, smoking status, Townsend deprivation index, MET and the muscle and fat IDPs including total muscle volume, thigh muscle volume, mid-thigh muscle volume, iliopsoas muscle volume, thigh IMAT volume, mid-thigh IMAT volume, mid-thigh IMAT/muscle volume and paraspinal muscle PDFF. Additionally, we incorporated paraspinal muscle iron concentration in our model due to its established association with adiposity and metabolic-related conditions (Moreno-Navarrete et al., 2016) We further applied an ordinal logistic regression model to investigate the association between muscle and fat IDPs on frailty given it has multiple ordered categories (not frail, pre-frail, and frail). Ordinal logistic regression models were adjusted for all the variables used in the above logistic regression models. The ordinal logistic regression models were performed using the R *MASS* package (Venables and Ripley, 2013). Because muscle and fat IDPs are associated with body composition measurements such as waist-to-hip ratio and BMI, adjustment for these categories might result in over-adjustment bias (Schisterman, Cole and Platt, 2009). Furthermore, HGS was excluded from logistic models since it is used in the definition of dynapenia. In all models iliopsoas muscle, thigh muscle, thigh IMAT volumes, and indices are shown as the sum of left and right volumes). All anthropometric variables and IDPs were standardised prior to inclusion in the logistic regression models.

Summaries of the logistic regression model are reported as odds ratios (OR) with 95% confidence intervals (CIs). To assess the performance of each logistic regression model we used the following metrics: Akaike Information Criterion (AIC), the area under the curve (AUC) of the receiver operating characteristic (ROC) with corresponding CIs for dynapenia and concordance index (c-index) for frailty. Lower AIC values indicate a better model fit, while higher AUC and c-index values suggest better discrimination between cases. AUC was calculated using the *pROC* R package (Robin et al., 2011), while the c-index was computed using the *rms* R package (Harrell, 2013). The Bonferroni-corrected threshold for statistical significance was *0*.*05*/ *583* = *8*.*6* × *10*^−*5*^.

## 3 Results

Typical images showing muscle and fat IDPs generated from each participant are shown in Figure 1. Image datasets were available from 44,520 individuals, with all required IDPs being successfully derived for each participant.

**Figure 1.**
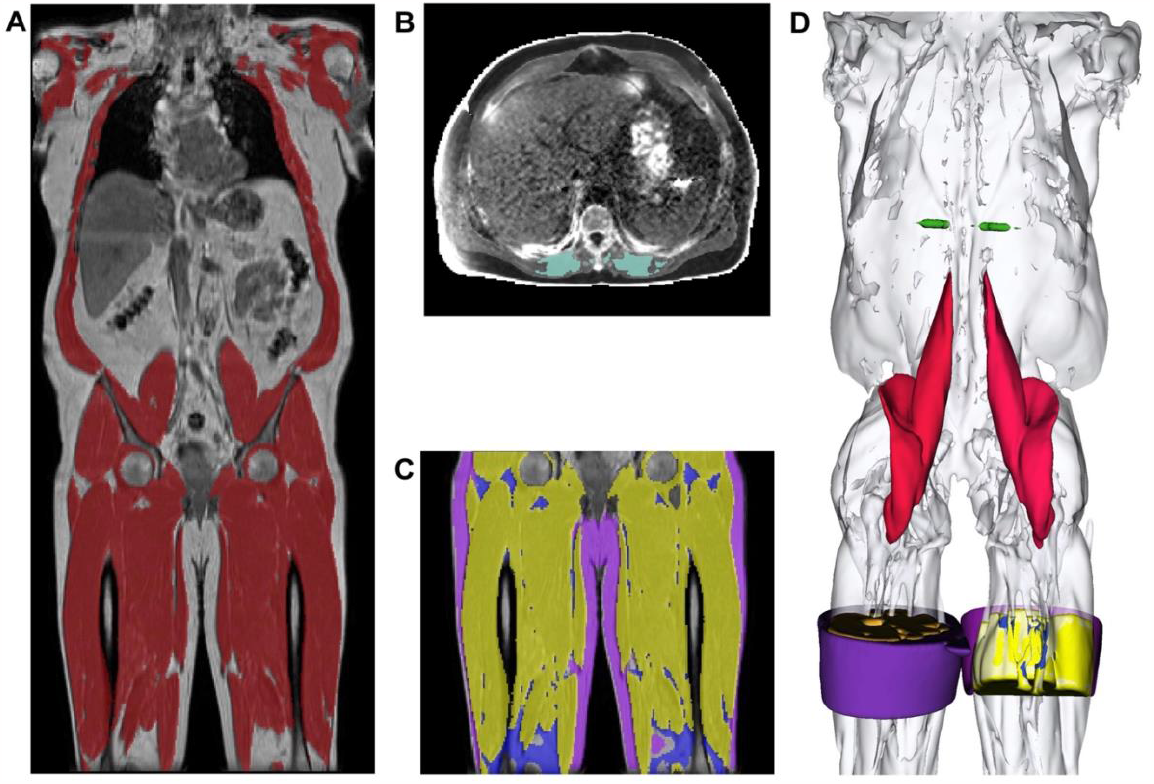
Segmentations from the UK Biobank abdominal MRI of a participant showing: **A)** the total muscle (dark red) overlaid on the coronal view of a neck-to-knee scan, **B)** the paraspinal muscle (green) in 2D quantitative MRI, **C)** the thigh muscles (yellow), thigh IMAT (blue), thigh SAT (purple) overlaid on the coronal view, and **D)** 3D renderings of the muscle segmentations for the following structures: mid-thigh muscle (yellow), mid-thigh IMAT (blue), mid-thigh subcutaneous adipose tissue (purple), paraspinal muscles (green), iliopsoas muscles (red) and total muscle (white).

### 3.1 Study population characteristics

Summary statistics for the study population are provided in Table 1, 51.7% were female participants with similar average age to male (female participants 63.54 ± 7.58 years and male participants 64.93 ± 7.83 years). The average BMI for female and male participants were 26.02 ± 4.72 kg/m^2^ (range 13.4 to 62.0 kg/m^2^) and 26.95 ± 3.90 kg/m^2^ (range 16.4 to 58.1 kg/m^2^) respectively. HGS in the dominant hand was 38.82 ± 8.85 kg in male participants and 23.98 ± 6.12 kg in female participants and the MET was reported as 24.03 ± 17.51 hours/week from female participants and 25.16 ± 17.47 hours/week from male participants (Table 1). The overall ethnic distribution for the cohort was 96.7% White, 1.1% Asian, 0.7% Black, 0.3% Chinese and 1% Others (see methods for details).

**Table 1.** Demographics of the participants (N=44,520), separated by sex.

|  | Full cohort | Female | Male | p-values |
| --- | --- | --- | --- | --- |
| N | 44,520 | 23,013 | 21,507 | - |
| White (N) | 43,059 | 22,277 | 20,782 | - |
| Asian (N) | 476 | 175 | 301 | - |
| Black (N) | 297 | 164 | 133 | - |
| Chinese (N) | 133 | 80 | 53 | - |
| Others (N) | 438 | 265 | 173 | - |
| Age (yrs.) | 64.21 ± 7.73 | 63.54 ± 7.58 | 64.93 ± 7.83 | $p < 10^{-16}$ |
| Weight (kg) | 75.99 ± 15.10 | 68.88 ± 13.03 | 83.60 ± 13.35 | $p < 10^{-16}$ |
| Height (cm) | 169.17 ± 9.27 | 162.72 ± 6.25 | 176.06 ± 6.64 | $p < 10^{-16}$ |
| BMI (kg/m<sup>2</sup>) | 26.47 ± 4.37 | 26.02 ± 4.72 | 26.95 ± 3.90 | $p < 10^{-16}$ |
| Waist Circumference (cm) | 88.37 ± 12.69 | 82.78 ± 11.83 | 94.34 ± 10.71 | $p < 10^{-16}$ |
| **Hip Circumference (cm)** | 100.76 ± 8.71 | 100.81 ± 9.83 | 100.70 ± 7.34 | $1.4 \times 10^{-9}$ |
| **Waist-to-Hip Ratio** | 0.88 ± 0.09 | 0.82 ± 0.07 | 0.94 ± 0.06 | $p < 10^{-16}$ |
| **Dominant HGS (kg)** | 31.17 ± 10.60 | 23.98 ± 6.12 | 38.82 ± 8.85 | $p < 10^{-16}$ |
| **Walking MET (hours/week)** | 9.16 ± 7.23 | 9.18 ± 7.33 | 9.15 ± 7.12 | 0.385 |
| **Moderate MET (hours/week)** | 8.55 ± 7.81 | 8.61 ± 7.88 | 8.49 ± 7.74 | 0.995 |
| **Vigorous MET (hours/week)** | 6.86 ± 7.94 | 6.25 ± 7.68 | 7.52 ± 8.16 | $p < 10^{-16}$ |
| **Total MET (hours/week)** | 24.58 ± 17.49 | 24.03 ± 17.51 | 25.16 ± 17.47 | $8.7 \times 10^{-13}$ |
| **Townsend deprivation index** | -1.88 ± 2.73 | -1.83 ± 2.74 | -1.93 ± 2.72 | $1.6 \times 10^{-6}$ |
| **Alcohol frequency (N)** |  |  |  |  |
| **Daily** | 7,585 | 3,118 | 4,467 | - |
| **1-4 times per week** | 24,014 | 11,802 | 12,212 | - |
| **1-3 times per month** | 5,130 | 3,081 | 2,049 | - |
| **Occasionally or never** | 7,469 | 4,841 | 2,628 | - |
| **Smoking status (N)** |  |  |  |  |
| **Never** | 27,481 | 15,035 | 12,446 | - |
| **Previous** | 15,067 | 7,068 | 7,999 | - |
| **Current** | 1,518 | 657 | 861 | - |
| **VAT (L)** | 3.95 ± 2.31 | 2.82 ± 1.57 | 5.16 ± 2.36 | $p < 10^{-16}$ |
| **ASAT (L)** | 8.47 ± 4.12 | 9.82 ± 4.37 | 7.01 ± 3.27 | $p < 10^{-16}$ |

Values are reported as mean and standard deviation for continuous variables and counts (N) for categorical variables. Significance refers to the p-value for a Wilcoxon rank-sums test, where the null hypothesis is the medians between the two groups (male and female participants) being equal. BMI: Body mass index; HGS: Hand grip strength; MET: Metabolic equivalents of task; VAT: Visceral adipose tissue; ASAT: Abdominal subcutaneous adipose tissue.

### 3.2 Muscle volume and quality characteristics

Total muscle in female and male participants were 14.04 ± 1.95 L and 21.80 ± 3.00 L (p < *8*.*6* × *10*^−*5*^) respectively, with a mean difference between the sexes of 7.76 ± 3.60 L, or 43% (Table 2). Similar proportional sex differences were observed for all other muscle IDPs. We also observed significant differences between left and right muscle volumes for the thigh, mid-thigh, and iliopsoas muscles in both female and male participants (Supplementary Table S2). Mid-thigh IMAT volumes, reflecting reduced muscle quality, were higher in male than female participants (55.19 ± 50.27 ml vs 48.19 ± 38.40 ml, respectively) (Table 2). However, this trend was reversed when IMAT was corrected for muscle volume in the mid-thigh, represented as mid-thigh IMAT/muscle ratio. In this context, female participants exhibited a percentage of 2.48 ± 1.86 %, whereas male participants demonstrated 1.97 ± 1.84 % (p < *8*.*6* × *10*^−*5*^) (Table 2). Paraspinal muscle PDFF, was 7.84 ± 4.03 % in female and 6.88 ± 3.64 % in male participants (p < *8*.*6* × *10*^−*5*^), while iron content showed a small but significant difference between sexes (Supplementary Table S2).

**Table 2.** Summary statistics for muscle volume and quality IDPs.

|  | **Full cohort**<br>(N = 44,520) | **Female**<br>(N = 23,013) | **Male**<br>(N = 21,507) | **p-values** |
| --- | --- | --- | --- | --- |
| **Muscle volume IDPs** |  |  |  |  |
| **Total Muscle Volume (L)** | 17.79 ± 4.62 | 14.04 ± 1.95 | 21.80 ± 3.00 | $p < 10^{-16}$ |
| **Total Muscle Volume Index (L/m<sup>2</sup>)** | 6.14 ± 1.12 | 5.30 ± 0.62 | 7.03 ± 0.81 | $p < 10^{-16}$ |
| **Thigh Muscle Volume (L)** | 8.45 ± 2.23 | 6.68 ± 1.01 | 10.34 ± 1.53 | $p < 10^{-16}$ |
| **Thigh Muscle Volume Index (L/m<sup>2</sup>)** | 2.91 ± 0.54 | 2.52 ± 0.31 | 3.33 ± 0.40 | $p < 10^{-16}$ |
| **Mid-thigh Muscle Volume (L)** | 2.28 ± 0.55 | 1.87 ± 0.28 | 2.73 ± 0.41 | $p < 10^{-16}$ |
| **Iliopsoas Muscle Volume (L)** | 0.64 ± 0.17 | 0.51 ± 0.08 | 0.78 ± 0.12 | $p < 10^{-16}$ |
| **Iliopsoas Muscle Volume Index (L/m<sup>2</sup>)** | 0.22 ± 0.04 | 0.19 ± 0.02 | 0.25 ± 0.03 | $p < 10^{-16}$ |
| **Muscle quality IDPs** |  |  |  |  |
| **Thigh IMAT Volume (L)** | 0.77 ± 0.34 | 0.71 ± 0.29 | 0.84 ± 0.37 | $p < 10^{-16}$ |
| **Thigh IMAT Volume Index (mL/m<sup>2</sup>)** | 269.82 ± 114.91 | 266.61 ± 109.33 | 273.24 ± 120.49 | 0.00039 |
| **Mid-thigh IMAT Volume (mL)** | 51.57 ± 44.67 | 48.19 ± 38.40 | 55.19 ± 50.27 | $p < 10^{-16}$ |
| **Mid-thigh IMAT/Muscle (%)** | 2.24 ± 1.87 | 2.48 ± 1.86 | 1.97 ± 1.84 | $p < 10^{-16}$ |
| **Paraspinal Muscle PDFF (%)** | 7.38 ± 3.88 | 7.84 ± 4.03 | 6.88 ± 3.64 | $p < 10^{-16}$ |
| **Paraspinal Muscle Iron Concentration (mg/g)** | 1.20 ± 0.12 | 1.20 ± 0.13 | 1.19 ± 0.10 | $p < 10^{-16}$ |

Values are reported as mean and standard deviation. Significance refers to the p-value for a Wilcoxon rank-sums test, where the null hypothesis is the medians between the two groups (male and female participants) being equal. IMAT: Intermuscular adipose tissue; PDFF: Proton density fat fraction.

### 3.3 Associations between dynapenia and frailty and muscle health

#### 3.3.1 Participant characteristics

From the overall cohort of 44,520 participants 3,261 (7.6%) were classified as having dynapenia, and 455 (1.1%) were classified as frail (Supplementary Table S3). Representative examples of MRI images from these subgroups are shown in Figure 2. The characteristics as well as the summaries of muscle and fat IDPs of dynapenia and frailty participants are presented in Supplementary Tables S4-S7. In summary, participants with and without dynapenia were aged 67.96 ± 7.17 and 63.86 ± 7.69 years old, respectively. Not-frail, pre-frail and frail participants were 63.65 ± 7.59, 64.94 ± 7.79 and 66.56 ± 8.04 years old, respectively. Participants with dynapenia also had lower physical activity levels and a higher waist-to-hip ratio (p < *8*.*6* × *10*^−*5*^). Similar results were observed for frailty. The average total muscle volume for participants without dynapenia was 17.92 ± 4.64 L whereas for participants with dynapenia was 16.49 ± 4.20 L (p < *8*.*6* × *10*^−*5*^). For participants without frailty and with pre-frailty, the average total muscle volume was 17.95 ± 4.67 and 17.70 ± 4.57 L, respectively, while frail participants had an average total muscle volume of 16.59 ± 4.13 L. Additionally, muscle quality measurements such as the mid-thigh IMAT/Muscle ratio was higher in participants with dynapenia and frailty (2.19 ± 1.84 for no dynapenia; 2.71 ± 2.17 % for dynapenia; 1.98 ± 1.59 for not-frail; 2.49 ± 1.93 for pre-frail and 3.73 ± 2.86 % for frail). Furthermore, there was a significant correlation between dominant HGS and muscle IDPs among participants without dynapenia for both male and female participants, showing the highest correlation with total muscle volume (male participants: ρ = 0.35; female participants ρ = 0.38, both *p* < *8*.*6* × *10*^−*5*^), however, this relationship lost significance in the presence of dynapenia (Supplementary Figure S1).

**Figure 2.**
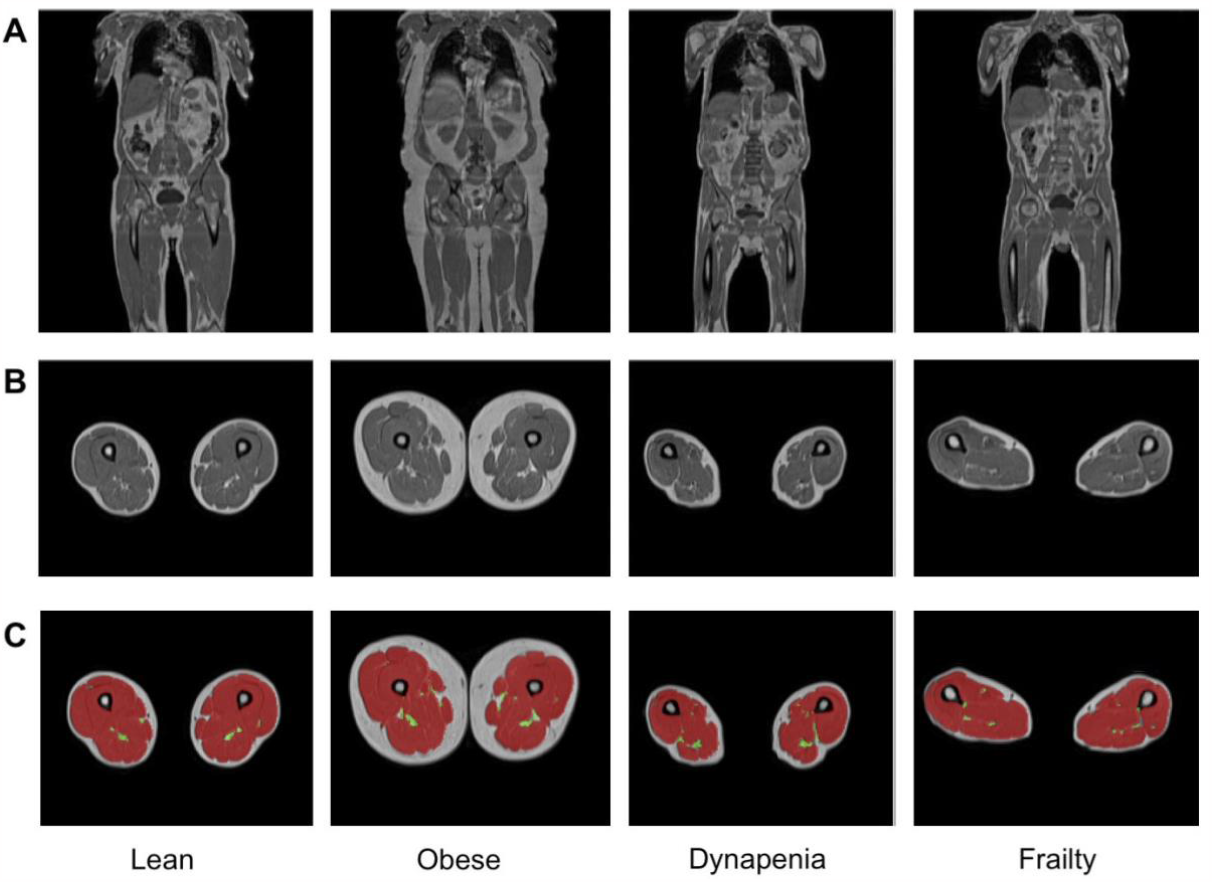
Representative samples of **A)** coronal and **B)** axial MRI views and **C)** the total muscle (red) and the thigh IMAT (green) segmentations overlaid on the axial views, obtained from lean, obese, dynapenic and frail participants from the Dixon acquisition.

#### 3.3.2 Logistic regression analysis

To determine which IDPs were associated with dynapenia and frailty, we created logistic regression models, adjusting for age, sex, ethnicity, alcohol intake frequency, smoking status, Townsend deprivation index and MET. Our findings reveal that both dynapenia and frailty were positively associated with age while displaying negative associations with alcohol and MET in all models. Dynapenia displayed a positive association with males only in models including muscle volume IDPs, while frailty was only positively associated in the models including total, thigh and iliopsoas muscle IDPs as well as mid-thigh IMAT/Muscle ratio and paraspinal muscle PDFF (Supplementary Tables S8, S9). Additionally, smaller muscle volume IDPs and larger muscle fat IDPs were associated with the presence of dynapenia and frailty. Specifically, dynapenia and frailty were negatively associated with total muscle volume (OR = 0.392 95% CI 0.361 - 0.426, *p* < *8*.*6* × *10*^−*5*^, for dynapenia; OR = 0.894 95% CI 0.854 - 0.935, *p* < *8*.*6* × *10*^−*5*^, for frailty). In contrast, thigh IMAT volume index demonstrated a positive association with increased odds ratios for dynapenia (OR = 1.129 95% CI 1.091 - 1.168, *p* < *8*.*6* × *10*^−*5*^) and frailty (OR = 1.410 95% CI 1.388 - 1.432, *p* < *8*.*6* × *10*^−*5*^). Paraspinal muscle PDFF was also significantly associated with dynapenia and frailty showing increased odds ratios (OR = 1.094 95% CI 1.059 - 1.130, *p* < *8*.*6* × *10*^−*5*^, for dynapenia; OR = 1.350 95% CI 1.327 - 1.372, *p* < *8*.*6* × *10*^−*5*^, for frailty) while, paraspinal muscle iron concentration was not significantly associated with any of these conditions.

#### 3.3.3 Performance evaluation

Performance judged by the model with the lowest AIC (Akaike’s Information Criterion) and the highest AUC and c-index (Supplementary Tables S8 - S10), showed that for dynapenia, total muscle (AUC = 0.707 (0.698, 0.716 95% CI)) and total thigh volume (AUC = 0.707 (0.697, 0.716 95% CI)) were the most informative IDPs (Supplementary Table S8), with muscle quality IDPs providing the least information. Interestingly, the indexed volumes demonstrated a relatively poor fit in their associations with dynapenia. Conversely, for frailty, measures of muscle quality including thigh IMAT volume index (c-index = 0.621) and mid-thigh IMAT/Muscle ratio (c-index = 0.618) were the most informative, with muscle volumes IDPs providing less information (Supplementary Tables S9, S10).

## 4 Discussion

In the present study we have utilised an automated deep-learning technique to accurately measure muscle volumes and quality in a large prospective cohort of middle-aged and older adults in the UK Biobank. We have confirmed and expanded on previous studies looking at the impact of age, sex, and lifestyle factors on muscle strength and have identified muscle volume and quality measurements being strongly associated with dynapenia and frailty, respectively (Kalyani, Corriere and Ferrucci, 2014). Specifically, male participants exhibited higher muscle volumes compared to females, while when considering mid-thigh IMAT/muscle ratio and paraspinal muscle PDFF, females displayed higher values. Additionally, the prevalence of dynapenia and frailty was relatively low, consistent with the UK Biobank’s generally healthy population. Both dynapenia and frailty were associated with increased age while they were negatively associated with alcohol and MET. Dynapenia was negatively associated with muscle volume and positively associated with mid-thigh IMAT/muscle and paraspinal PDFF levels. Muscle volume IDPs including total muscle and thigh measurements were more informative for dynapenia, while measures of muscle quality, specifically thigh IMAT volume index and mid-thigh IMAT/muscle ratio, were more informative for frailty, suggesting that different measurements may have distinct diagnostic values for various conditions.

Muscle volumes were significantly higher in male compared with female participants, even after correcting for height. We further show that total thigh IMAT content was significantly higher in male compared to female participants, however when presented as the mid-thigh IMAT/muscle ratio, this relationship was reversed, with higher levels of mid-thigh IMAT/muscle found in female participants. Similarly, paraspinal muscle PDFF, was also significantly higher in female participants. This clearly highlights the impact of the type of measurement on possible outcomes. Previously studies have reported higher whole-body and thigh IMAT levels in men (Gallagher et al., 2005; Sparks, Goodpaster and Bergman, 2021), but these were generally not corrected for muscle volume. Although, correcting for muscle mass negated reported sex differences in IMAT (Kim et al., 2017).

The prevalence of dynapenia (7.6%) and frailty (1.1%) were relatively low in our cohort, in agreement with reports that the UK Biobank cohort is a comparatively healthy population (Lyall et al., 2022). In this study, we defined dynapenia based on low muscle strength (Dodds et al., 2020; Wilkinson et al., 2022), without including reduced muscle mass/quality and/or reduced physical performance. Dynapenia was positively associated in men only in the model incorporating muscle volume measurements, while frailty showed a positive association in models that included total muscle, thigh muscle, iliopsoas muscle, mid-thigh IMAT/Muscle ratio and paraspinal muscle PDFF IDPs. Additionally, participants with both dynapenia and frailty demonstrated positive associations with age and lower physical activity level in all models, confirming earlier literature reports (Bouchard, Dionne and Brochu, 2009).

Our study further aimed to identify which of our measured muscle IDPs were associated with these health conditions, and which would potentially provide the best prognostic tool for identifying this population. We found that dynapenia was associated with reduced muscle volume and increased levels of mid-thigh IMAT/muscle and paraspinal PDFF. Notably, for dynapenia, the most informative muscle volume measurements were those with a greater proportion of body included such as total muscle or the entire thigh. On the other hand, for frailty markers of muscle quality, particularly thigh IMAT volume index to height squared and mid-thigh IMAT/Muscle were the most informative IDPs. This suggests that different IDPs may have different diagnostic values for different conditions. Interestingly, we observed that the muscle volumes indexed to height squared demonstrated a relatively poor fit in their associations with dynapenia.

A previous observational study, investigating the feasibility of using psoas or paraspinal muscles at the same axial slice as well as mid-thigh measurement to predict sarcopenia, found that the psoas muscle was more accurate than the paraspinal muscle, although inferior to mid-thigh measurement (Morrell et al., 2016). Additional studies in the UK biobank investigating associations between low muscle mass, malnutrition and sarcopenia with cancer (Kiss et al., 2023), found that in participants with obesity, only the BMI-adjusted muscle mass identified cases of low muscle mass and sarcopenia. They also reported that BMI-adjusted methods may be more suitable for determining these cases of malnutrition and sarcopenia; however, height-adjusted muscle mass showed a higher risk of these conditions occurring. Furthermore, in a previous study, Linge et al, (Linge, Heymsfield and Dahlqvist Leinhard, 2020) reported that body size adjustments led to unnormalized correlations between muscle volume and body size. However, they also report that adjustments considering the distribution of height-adjusted measurements for each body size value significantly strengthened the associations between muscle volume and both functional and health outcomes. Future work using longitudinal data may provide better insights into which muscle volume and quality measurement can best associate with or even predict these health conditions.

Our study is not without limitations. The UK Biobank is a large cross-sectional study that is subject to selection bias with a “healthier” cohort than the wider UK population, excludes younger participants and potentially more severe clinical cases (Munafò *et al*., 2018). Furthermore, as the UK Biobank study includes a predominantly White population, future work is needed to further investigate imbalanced population data. Another potential limitation of this study is that although food intake such as energy and protein intake is widely used to assess the loss of muscle mass and function (Celis-Morales *et al*., 2018; Gedmantaite et al., 2020), these dietary parameters were not used in this study as they are reported 6.8 ± 1.5 years prior to the UK Biobank first imaging visit. Also, given the UK Biobank neck-to-knee acquisition, measurement of whole-body muscle volume could not be achieved, thus limiting direct comparisons with some literature values (Karlsson *et al*., 2015). We estimate that the current protocol lacks c.a. 9.75% of total muscle, corresponding to both arms and lower legs. However, as our findings were robust to the selection of muscle volume or quality measure, we consider it unlikely that results with a whole body measure would be dramatically different.

## 5 Conclusions

We present a fully automated method for accurately measuring muscle volumes and quality, making it suitable for applications in large-scale population-based studies. Our investigation revealed notable sex-related variations, with male participants exhibiting higher muscle volumes. In contrast, female participants displayed more significant mid-thigh IMAT to muscle ratio and paraspinal muscle PDFF. We further explored how the choice of specific muscle and fat measurements can impact the identification of diseases. Total muscle volume proved to be the most informative parameter for identifying dynapenia, whereas for frailty, muscle quality, especially thigh IMAT volume indexed to height squared, showed the strongest associations. Our research also highlights that the choice of muscle measurements can significantly affect the assessment of disease. However, it is essential to note that most of these measurements consistently reveal differences related to age, sex, and clinical conditions, emphasising their value in assessing muscular health. Our findings underscore the significance of these muscle metrics in evaluating the impact of various factors on muscular health within diverse populations. These metrics can potentially serve as valuable tools for future research and clinical applications.

## 7 Ethics Statement

The data resources used in this study have approval from ethics committees. Full anonymised images and participants metadata from the UK Biobank cohort was obtained through UK Biobank Access Application number 44584. The UK Biobank has approval from the North West Multi-Centre Research Ethics Committee (REC reference: 11/NW/0382), and obtained written informed consent from all participants prior to the study. All methods were performed in accordance with the relevant guidelines and regulations as presented by the relevant authorities, including the Declaration of Helsinki https://www.ukbiobank.ac.uk/learn-more-about-uk-biobank/about-us/ethics.

## 8 Data Availability Statement

The UK Biobank resource is available to bona fide researchers for health-related research in the public interest. All researchers who wish to access the research resource must register with UK Biobank by completing the registration form in the Access Management System (AMS - https://bbams.ndph.ox.ac.uk/ams/).

## 10 Author Contributions

J.D.B., E.L.T., M.T. and M.C. conceived the study. J.D.B., B.W., E.L.T., N.B. and M.T. designed the study. M.T., N.B., B.W., Y.L., E.P.S. and M.C. implemented the methods and performed the data analysis. M.T. defined the disease categories. M.T. performed the statistical analysis. E.L.T., B.W., M.T., J.D.B., R.S. and N.B. drafted the manuscript. All authors read and approved the manuscript.

## 11 Acknowledgements

This research has been conducted using the UK Biobank Resource under Application Number 44584.

## 12 Supplementary Material

**Supplementary Figure S1**

Scatterplot showing the correlation coefficients between muscle volume IDPs and dominant hand grip strength, separated by presence of dynapenia (blue for TRUE, red for FALSE) in both genders. *indicate statistically significant for *p* < *0*.*05*, **indicate statistically significant after Bonferroni correction (*p* = *8*.*6* × *10*^−*5*^).

## References

Bouchard, D.R., Dionne, I.J. and Brochu, M. (2009) ‘Sarcopenic/Obesity and Physical Capacity in Older Men and Women: Data From the Nutrition as a Determinant of Successful Aging (NuAge)—the Quebec Longitudinal Study’, Obesity, 17(11), p. 2082–2088.

Bradbury, K.E. et al. (2017) ‘Association between physical activity and body fat percentage, with adjustment for BMI: a large cross-sectional analysis of UK Biobank’, BMJ open, 7(3), p. e011843.

Celis-Morales, C.A. et al. (2018) ‘Associations of Dietary Protein Intake With Fat-Free Mass and Grip Strength: A Cross-Sectional Study in 146,816 UK Biobank Participants’, American journal of epidemiology, 187(11), p. 2405–2414.

Cruz-Jentoft, A.J. et al. (2019) ‘Sarcopenia: revised European consensus on definition and diagnosis’, Age and ageing, 48(1), p. 16–31.

Dodds, R.M. et al. (2014) ‘Grip strength across the life course: normative data from twelve British studies’, PloS one, 9(12), p. e113637.

Dodds, R.M. et al. (2020) ‘Sarcopenia, long-term conditions, and multimorbidity: findings from UK Biobank participants’, Journal of cachexia, sarcopenia and muscle, 11(1), p. 62–68.

Ebbeling, L. et al. (2014) ‘Psoas:lumbar vertebra index: central sarcopenia independently predicts morbidity in elderly trauma patients’, European journal of trauma and emergency surgery: official publication of the European Trauma Society, 40(1), p. 57–65.

Fitzpatrick, J.A. et al. (2020) ‘Large-scale analysis of iliopsoas muscle volumes in the UK Biobank’, Scientific reports, 10(1), p. 20215.

Gallagher, D. et al. (2005) ‘Adipose tissue in muscle: a novel depot similar in size to visceral adipose tissue’, The American journal of clinical nutrition, 81(4), p. 903–910.

Gedmantaite A. et al. (2020) ‘Associations between diet and handgrip strength: a crosssectional study from UK Biobank’ Mechanisms of ageing and development, 189, p. 111269.

Hanlon, P. et al. (2018) ‘Frailty and pre-frailty in middle-aged and older adults and its association with multimorbidity and mortality: a prospective analysis of 493 737 UK Biobank participants’, The Lancet. Public health, 3(7), p. e323–e332.

Harrell, F.E. (2013) Regression Modeling Strategies: With Applications to Linear Models, Logistic Regression, and Survival Analysis. Springer Science & Business Media.

Hofsteenge, G.H., Chinapaw, M.J.M. and Weijs, P.J.M. (2015) ‘Fat-free mass prediction equations for bioelectric impedance analysis compared to dual energy X-ray absorptiometry in obese adolescents: a validation study’, BMC pediatrics, 15, p. 158.

Jones, G. et al. (2021) ‘Genome-wide meta-analysis of muscle weakness identifies 15 susceptibility loci in older men and women’, Nature communications, 12(1), p. 654.

Kalyani, R.R., Corriere, M. and Ferrucci, L. (2014) ‘Age-related and disease-related muscle loss: the effect of diabetes, obesity, and other diseases’, The lancet. Diabetes & endocrinology, 2(10), p. 819–829.

Karlsson, A. et al. (2015) ‘Automatic and quantitative assessment of regional muscle volume by multi-atlas segmentation using whole-body water-fat MRI’, Journal of magnetic resonance imaging: JMRI, 41(6), p. 1558–1569.

Kasahara, R. et al. (2017) ‘A Low Psoas Muscle Index before Treatment Can Predict a Poorer Prognosis in Advanced Bladder Cancer Patients Who Receive Gemcitabine and Nedaplatin Therapy’, BioMed research international, 2017, p. 7981549.

Kim, J.C. et al. (2019) ‘Natural aging course of paraspinal muscle and back extensor strength in community-dwelling older adults (sarcopenia of spine, SarcoSpine): a prospective cohort study protocol’, BMJ open, 9(9), p. e032443.

Kim, J.E. et al. (2017) ‘Intermuscular Adipose Tissue Content and Intramyocellular Lipid Fatty Acid Saturation Are Associated with Glucose Homeostasis in Middle-Aged and Older Adults’, Endocrinology and metabolism (Seoul, Korea), 32(2), p. 257–264.

Kiss, N. et al. (2023) ‘Low muscle mass, malnutrition, sarcopenia, and associations with survival in adults with cancer in the UK Biobank cohort’, Journal of cachexia, sarcopenia and muscle [Preprint]. Available at: 10.1002/jcsm.13256.

Linge, J. et al. (2018) ‘Body Composition Profiling in the UK Biobank Imaging Study’, Obesity, 26(11), p. 1785–1795.

Linge, J., Ekstedt, M. and Dahlqvist Leinhard, O. (2021) ‘Adverse muscle composition is linked to poor functional performance and metabolic comorbidities in NAFLD’, JHEP reports : innovation in hepatology, 3(1), p. 100197.

Linge, J., Heymsfield, S.B. and Dahlqvist Leinhard, O. (2020) ‘On the Definition of Sarcopenia in the Presence of Aging and Obesity-Initial Results from UK Biobank’, The journals of gerontology. Series A, Biological sciences and medical sciences, 75(7), p. 1309–1316.

Littlejohns, T.J. et al. (2020) ‘The UK Biobank imaging enhancement of 100,000 participants: rationale, data collection, management and future directions’, Nature communications, 11(1), p. 2624.

Liu, Y. et al. (2021) ‘Genetic architecture of 11 organ traits derived from abdominal MRI using deep learning’, eLife, 10. Available at: 10.7554/eLife.65554.

Lyall, D.M. et al. (2022) ‘Quantifying bias in psychological and physical health in the UK Biobank imaging sub-sample’, Brain Communications, 4(3), p. fcac119.

Masaki, M. et al. (2016) ‘Association of walking speed with sagittal spinal alignment, muscle thickness, and echo intensity of lumbar back muscles in middle-aged and elderly women’, Aging clinical and experimental research, 28(3), p. 429–434.

Mitchell, W.K. et al. (2012) ‘Sarcopenia, dynapenia, and the impact of advancing age on human skeletal muscle size and strength; a quantitative review’, Frontiers in physiology, 3, p. 260.

Moreno-Navarrete, J.M. et al. (2016) ‘Obesity Is Associated With Gene Expression and Imaging Markers of Iron Accumulation in Skeletal Muscle’, The Journal of clinical endocrinology and metabolism, 101(3), p. 1282–1289.

Morrell, G.R. et al. (2016) ‘Psoas Muscle Cross-sectional Area as a Measure of Whole-body Lean Muscle Mass in Maintenance Hemodialysis Patients’, Journal of renal nutrition: the official journal of the Council on Renal Nutrition of the National Kidney Foundation, 26(4), pp. 258–264.

Munafò, M.R. et al. (2018) ‘Collider scope: when selection bias can substantially influence observed associations’, International journal of epidemiology, 47(1), p. 226–235.

O’Donnell, J. et al. (2020) ‘Self-reported and objectively measured physical activity in people with and without chronic heart failure: UK Biobank analysis’, Open heart, 7(1), p. e001099.

Rantanen, T. et al. (1999) ‘Midlife hand grip strength as a predictor of old age disability’, JAMA: the journal of the American Medical Association, 281(6), p. 558–560.

Robin, X. et al. (2011) ‘pROC: an open-source package for R and S+ to analyze and compare ROC curves’, BMC bioinformatics, 12(1), p. 1–8.

Scafoglieri, A. and Clarys, J.P. (2018) ‘Dual energy X-ray absorptiometry: gold standard for muscle mass?’, Journal of cachexia, sarcopenia and muscle, pp. 786–787.

Schisterman, E.F., Cole, S.R. and Platt, R.W. (2009) ‘Overadjustment bias and unnecessary adjustment in epidemiologic studies’, Epidemiology, 20(4), p. 488–495.

Schweitzer, L. et al. (2015) ‘What is the best reference site for a single MRI slice to assess whole-body skeletal muscle and adipose tissue volumes in healthy adults?’, The American journal of clinical nutrition, 102(1), p. 58–65.

Silva R.R. et al. (2022) ‘Dynapenia in all-cause mortality and its relationship with sedentary behavior in community-dwelling older adults’, Sports Medicine and Health Science, 4(4), p. 253–259.

Sizoo, D. et al. (2021) ‘Measuring Muscle Mass and Strength in Obesity: a Review of Various Methods’, Obesity surgery, 31(1), p. 384–393.

Sparks, L.M., Goodpaster, B.H. and Bergman, B.C. (2021) ‘The Metabolic Significance of Intermuscular Adipose Tissue: Is IMAT a Friend or a Foe to Metabolic Health?’, Diabetes, 70(11), pp. 2457–2467.

Ungar, A. and Marchionni, N. (2017) Cardiac Management in the Frail Elderly Patient and the Oldest Old. Springer.

Venables, W.N. and Ripley, B.D. (2013) Modern Applied Statistics with S-PLUS. Springer Science & Business Media.

Wilkinson, L. (2011) ‘ggplot2: Elegant Graphics for Data Analysis by WICKHAM, H’, Biometrics, pp. 678–679. Available at: 10.1111/j.1541-0420.2011.01616.x.

Wilkinson, T.J. et al. (2022) ‘Sarcopenic obesity and the risk of hospitalization or death from coronavirus disease 2019: findings from UK Biobank’, JCSM rapid communications, 5(1), pp. 3–9.

Wood, J.C. et al. (2005) ‘MRI R2 and R2* mapping accurately estimates hepatic iron concentration in transfusion-dependent thalassemia and sickle cell disease patients’, Blood, 106(4), pp. 1460–1465.

Zillikens, M.C. et al. (2017) ‘Large meta-analysis of genome-wide association studies identifies five loci for lean body mass’, Nature communications, 8(1), p. 80.

