## Supplementary_Material for "Precision MRI phenotyping of muscle volume and quality at a population scale"

#### Supplementary Tables

**Supplementary Table S1.** Demographics and frailty indicators for the participants (N= 44,520), separated by gender.

| | **Full cohort** (N = 44,520) | **Female** (N = 23,013) | **Male** (N = 21,507) |
|---|---|---|---|
| **Weight loss (N)** | 7,840 | 3,873 | 3,967 |
| **Exhaustion (N)** | 3,467 | 2,146 | 1,321 |
| **Physical activity (N)** | | | |
| **Low physical activity** | 150 | 98 | 52 |
| **Activity less than once a week** | 12,925 | 6,859 | 6,066 |
| **Walking pace (N)** | | | |
| **Slow** | 2,007 | 1,095 | 912 |
| **Steady** | 21,275 | 10,838 | 10,437 |
| **Brisk** | 19,346 | 10,049 | 9,297 |
| **Low grip strength* (N)** | 7,712 | 3,897 | 3,815 |

Values are reported as counts (N). * Definition used in original description by (Fried *et al.*, 2001), stratified by sex and body mass index.

**Supplementary Table S2.** Summary statistics for skeletal muscle and fat IDPs. Values are reported as mean and standard deviation.

| | **Full cohort** (N = 44,520) | **Female** (N = 23,013) | **Male** (N = 21,507) |
|---|---|---|---|
| **Muscle volume IDPs** | | | |
| **Left Thigh Muscle Volume (L)** | 4.22 ± 1.12 | 3.34 ± 0.51 | 5.16 ± 0.77** |
| **Right Thigh Muscle Volume (L)** | 4.23 ± 1.13 | 3.35 ± 0.51 | 5.18 ± 0.78** |
| **L-R Thigh Muscle Volume Difference (L)** | -0.01 ± 0.22 | -0.004 ± 0.17 | -0.02 ± 0.27** |
| **Left Mid-thigh Muscle Volume (L)** | 1.13 ± 0.27 | 0.93 ± 0.14 | 1.35 ± 0.21** |
| **Right Mid-thigh Muscle Volume (L)** | 1.15 ± 0.28 | 0.94 ± 0.14 | 1.38 ± 0.21** |
| **L-R Mid-thigh Muscle Volume Difference (L)** | -0.02 ± 0.07 | -0.01 ± 0.06 | -0.03 ± 0.08** |
| **Left Iliopsoas Muscle Volume (L)** | 0.31 ± 0.09 | 0.25 ± 0.04 | 0.39 ± 0.06** |

| | | | |
|---|---|---|---|
| **Right Iliopsoas Muscle Volume (L)** | 0.33 ± 0.09 | 0.26 ± 0.04 | 0.40 ± 0.06** |
| **L-R Iliopsoas Muscle Volume Difference (L)** | -0.01 ± 0.02 | -0.01 ± 0.02 | -0.01 ± 0.02* |
| **Muscle quality IDPs** | | | |
| **Left Thigh IMAT Volume (L)** | 0.39 ± 0.17 | 0.36 ± 0.15 | 0.42 ± 0.19** |
| **Right Thigh IMAT Volume (L)** | 0.39 ± 0.17 | 0.35 ± 0.15 | 0.42 ± 0.19** |
| **L-R Thigh IMAT Volume Difference (L)** | 0.00031 ± 0.05 | 0.0041 ± 0.04 | -0.0038 ± 0.05** |
| **Left Mid-thigh IMAT Volume (mL)** | 25.69 ± 22.57 | 24.16 ± 18.95 | 27.32 ± 25.79** |
| **Right Mid-thigh IMAT Volume (mL)** | 25.88 ± 24.11 | 24.03 ± 20.78 | 27.87 ± 27.10** |
| **L-R Mid-thigh IMAT Volume Difference (mL)** | -0.19 ± 13.64 | 0.13 ± 10.35 | -0.54 ± 16.45** |
| **Thigh SAT Volume (L)** | 7.48 ± 3.03 | 8.98 ± 2.98 | 5.87 ± 2.14** |
| **Thigh SAT Volume Index (L/m²)** | 2.67 ± 1.20 | 3.39 ± 1.11 | 1.89 ± 0.68** |
| **Left Thigh SAT Volume (L)** | 3.74 ± 1.52 | 4.48 ± 1.50 | 2.94 ± 1.08** |
| **Right Thigh SAT Volume (L)** | 3.74 ± 1.55 | 4.51 ± 1.53 | 2.93 ± 1.08** |
| **L-R Thigh SAT Volume Difference (L)** | -0.0089 ± 0.44 | -0.03 ± 0.53 | 0.01 ± 0.31** |
| **Mid-thigh SAT Volume (L)** | 1.67 ± 0.75 | 2.07 ± 0.76 | 1.25 ± 0.46** |
| **Left Mid-thigh SAT Volume (ml)** | 0.83 ± 0.37 | 1.02 ± 0.38 | 0.63 ± 0.23** |
| **Right Mid-thigh SAT Volume (L)** | 0.84 ± 0.39 | 1.04 ± 0.39 | 0.62 ± 0.24** |
| **L-R Mid-thigh SAT Volume Difference (L)** | -0.0063 ± 0.10 | -0.02 ± 0.12 | 0.007 ± 0.08** |

Significance refers to the p-value for a Wilcoxon rank-sums test, where the null hypothesis is the medians between the two groups (male and female participants) being equal. ** indicate statistically significant after Bonferroni correction ($p = 8.6 \times 10^{-5}$). IMAT: Intermuscular adipose tissue; SAT: Subcutaneous adipose tissue.

**Supplementary Table S3.** Summary of the participants defined with dynapenia and frailty.

| | **Full cohort** (N = 44,520) | **Female** (N = 23,013) | **Male** (N = 21,507) |
|---|---|---|---|
| **Dynapenia** | 3,261 (7.6%) | 1,668 (7.6%) | 1,593 (7.7%) |
| **Frail** | 455 (1.1%) | 273 (1.3%) | 182 (0.90%) |
| **Pre-Frail** | 15,686 (37.8%) | 7,983 (37.5%) | 7,703 (38.2%) |
| **Not-Frail** | 25,349 (61.1%) | 12,056 (61.3%) | 12,293 (60.9%) |

Note that there are overall 1,658 (914F/744M) missing data for dynapenia, and 3,030 (1,701F/1,329M) missing data for frailty.

**Supplementary Table S4.** Demographics of the participants with no dynapenia (N=39,601) and with dynapenia (N=3,261).

| | **No dynapenia** (N = 39,601) | **Dynapenia** (N = 3,261) |
|---|---|---|
| **Sex** | 19,170M / 20,431F | 1,593M / 1,668F |
| **White (N)** | 38,349 | 3,130 |
| **Asian (N)** | 392 | 67 |
| **Black (N)** | 270 | 9 |
| **Chinese (N)** | 112 | 13 |
| **Others (N)** | 390 | 32 |
| **Age (yrs.)** | 63.86 ± 7.69 | 67.96 ± 7.17** |
| **Weight (kg)** | 76.22 ± 15.10 | 74.26 ± 15.08** |
| **Height (cm)** | 169.40 ± 9.25 | 166.21 ± 9.05** |
| **BMI (kg/m²)** | 26.47 ± 4.35 | 26.80 ± 4.64* |
| **Waist circumference (cm)** | 88.26 ± 12.64 | 89.59 ± 13.27** |
| **Hip circumference (cm)** | 100.76 ± 8.67 | 100.60 ± 9.14 |
| **Waist-to-Hip Ratio** | 0.87 ± 0.09 | 0.89 ± 0.09** |
| **Dominant HGS (kg)** | 32.33 ± 10.04 | 17.09 ± 6.11** |
| **Walking MET (hours/week)** | 9.57 ± 7.14 | 8.70 ± 7.07** |
| **Moderate MET (hours/week)** | 8.92 ± 7.77 | 8.25 ± 7.91** |
| **Vigorous MET (hours/week)** | 7.26 ± 8.00 | 5.43 ± 7.41** |
| **Total MET (hours/week)** | 25.74 ± 17.14 | 22.38 ± 17.01** |
| **Townsend deprivation index** | -1.90 ± 2.71 | -1.80 ± 2.79 |
| **Alcohol frequency (N)** | | |
| **Daily** | 6,802 | 534 |
| **1-4 times per week** | 21,548 | 1,615 |
| **1-3 times per months** | 4,552 | 352 |
| **Occasional or never** | 6,468 | 736 |
| **Smoking status (N)** | | |
| **Never** | 24,540 | 1,994 |
| **Previous** | 13,352 | 1,124 |

| | | |
|---|---|---|
| Current | 1,372 | 99 |
| **VAT (L)** | 3.93 ± 2.31 | 4.23 ± 2.36** |
| **ASAT (L)** | 8.44 ± 4.12 | 8.84 ± 4.20** |

Values are reported as mean and standard deviation for continuous variables and counts (N) for categorical variables. Significance refers to the p-value for a Wilcoxon rank-sums test, where the null hypothesis is the medians between the two groups (no dynapenia and dynapenia participants) being equal. * indicate statistically significant for $p < 0.05$, ** indicate statistically significant after Bonferroni correction ($p = 8.6 \times 10^{-5}$). Note that there are overall 1,658 (914F/744M) missing data for dynapenia. M: Male; F: Female; BMI: Body mass index; HGS: Hand grip strength; MET: Metabolic equivalents of task; VAT: Visceral adipose tissue; ASAT: Abdominal subcutaneous adipose tissue.

**Supplementary Table S5.** Demographics of the participants with not-frail (N = 25,349), pre-frail (N = 15,686) and with frail (N=455).

| | **Not Frail** (N = 25,349) | **Pre Frail** (N = 15,686) | **Frail** (N = 455) |
|---|---|---|---|
| **Sex** | 12,293M/13,056F | 7,703M/7,983F | 182M/273F |
| **White (N)** | 24,666 | 15,102 | 431 |
| **Asian (N)** | 200 | 219 | 7 |
| **Black (N)** | 135 | 108 | 5 |
| **Chinese (N)** | 69 | 52 | 1 |
| **Others (N)** | 222 | 168 | 9 |
| **Age (yrs.)** | 63.65 ± 7.59 | 64.94 ± 7.79 | 66.56 ± 8.04 |
| **Weight (kg)** | 74.74 ± 14.45 | 77.55 ± 15.42 | 82.77 ± 17.89 |
| **Height (cm)** | 169.81 ± 9.18 | 168.39 ± 9.32 | 165.83 ± 8.91 |
| **BMI (kg/m²)** | 25.81 ± 3.90 | 27.27 ± 4.60 | 30.05 ± 5.83 |
| **Waist circumference (cm)** | 86.72 ± 12.02 | 90.22 ± 12.90 | 96.58 ± 14.84 |
| **Hip circumference (cm)** | 99.72 ± 7.92 | 101.83 ± 9.18 | 106.46 ± 12.16 |
| **Waist-to-Hip Ratio** | 0.87 ± 0.09 | 0.89 ± 0.09 | 0.91 ± 0.09 |
| **Dominant HGS (kg)** | 33.47 ± 9.95 | 28.08 ± 10.62 | 20.53 ± 8.46 |
| **Walking MET (hours/week)** | 9.95 ± 7.07 | 9.36 ± 7.13 | 7.08 ± 7.00 |
| **Moderate MET (hours/week)** | 9.32 ± 7.74 | 8.75 ± 7.77 | 6.66 ± 7.47 |
| **Vigorous MET (hours/week)** | 7.78 ± 8.05 | 6.71 ± 7.89 | 3.41 ± 6.33 |
| **Total MET (hours/week)** | 27.05 ± 16.92 | 24.81 ± 16.96 | 17.15 ± 15.31 |
| **Townsend deprivation index** | -2.00 ± 2.67 | -1.83 ± 2.74 | -1.18 ± 3.06 |
| **Alcohol frequency (N)** | | | |

| | | | |
|---|---|---|---|
| **Daily** | 4,631 | 2,490 | 57 |
| **1-4 times per week** | 14,468 | 8,112 | 172 |
| **1-3 times per months** | 2,735 | 1,925 | 59 |
| **Occasional or never** | 3,506 | 3,153 | 167 |
| **Smoking status (N)** | | | |
| **Never** | 16,175 | 9,496 | 257 |
| **Previous** | 8,315 | 5,567 | 176 |
| **Current** | 789 | 558 | 19 |
| **VAT (L)** | 3.70 ± 2.23 | 4.24 ± 2.34 | 5.10 ± 2.63 |
| **ASAT (L)** | 7.89 ± 3.71 | 9.10 ± 4.39 | 11.67 ± 5.45 |

Values are reported as mean and standard deviation for continuous variables and counts (N) for categorical variables. One-way ANOVA was used to compare the following groups: not frail - pre frail, not frail - frail and pre frail - frail participants. After Bonferroni correction ($p = 8.6 \times 10^{-5}$), all group comparisons of the continuous variables were statistically significant. Note that there are overall 3,030 (1,701F/1,329M) missing data for frailty. M: Male; F: Female; BMI: Body mass index; HGS: Hand grip strength; MET: Metabolic equivalents of task; VAT: Visceral adipose tissue; ASAT: Abdominal subcutaneous adipose tissue.

**Supplementary Table S6.** Summary statistics for muscle volume and quality IDPs of the participants with no dynapenia (N=39,601) and with dynapenia (N=3,261).

| | **No dynapenia** (N = 39,601) | **dynapenia** (N = 3,261) |
|---|---|---|
| **Muscle volume IDPs** | | |
| **Total Muscle Volume (L)** | 17.92 ± 4.64 | 16.49 ± 4.20** |
| **Total Muscle Volume Index (L/m²)** | 6.16 ± 1.12 | 5.89 ± 1.07** |
| **Thigh Muscle Volume (L)** | 8.51 ± 2.24 | 7.80 ± 2.03** |
| **Thigh Muscle Volume Index (L/m²)** | 2.93 ± 0.54 | 2.79 ± 0.52** |
| **Mid-thigh Muscle Volume (L)** | 2.30 ± 0.55 | 2.12 ± 0.50** |
| **Iliopsoas Muscle Volume (L)** | 0.65 ± 0.17 | 0.59 ± 0.15** |
| **Iliopsoas Muscle Volume Index (L/m²)** | 0.22 ± 0.04 | 0.21 ± 0.04** |
| **Muscle quality IDPs** | | |
| **Thigh IMAT Volume (L)** | 0.77 ± 0.34 | 0.82 ± 0.38** |
| **Thigh IMAT Volume Index (mL/m²)** | 267.52 ± 113.36 | 297.08 ± 128.94** |
| **Mid-thigh IMAT Volume (mL)** | 50.97 ± 44.19 | 59.11 ± 49.75** |
| **Mid-thigh IMAT/Muscle (%)** | 2.19 ± 1.84 | 2.71 ± 2.17** |

| | | |
|---|---|---|
| **Paraspinal Muscle PDFF (%)** | 7.28 ± 3.79 | 8.35 ± 4.50** |
| **Paraspinal Muscle Iron Concentration (mg/g)** | 1.20 ± 0.12 | 1.20 ± 0.12 |

Values are reported as mean and standard deviation.Significance refers to the p-value for a Wilcoxon rank-sums test, where the null hypothesis is the medians between the two groups (no dynapenia and dynapenia participants) being equal. ** indicate statistically significant after Bonferroni correction ($p = 8.6 \times 10^{-5}$). IMAT: Intermuscular adipose tissue; PDFF: Proton density fat fraction.

**Supplementary Table S7.** Summary statistics for muscle volume and quality IDPs of the participants with not-frail (N = 25,349), pre-frail (N = 15,686) and with frail (N=455).

| | **Not Frail**[1] (N = 25,349) | **Pre Frail**[2] (N = 15,686) | **Frail**[3] (N = 455) |
|---|---|---|---|
| **Muscle volume IDPs** | | | |
| **Total Muscle Volume (L)** | 17.95 ± 4.67[2,3] | 17.70 ± 4.57[1,3] | 16.59 ± 4.13[1,2] |
| **Total Muscle Volume Index (L/m²)** | 6.14 ± 1.12 | 6.16 ± 1.12 | 5.96 ± 1.08 |
| **Thigh Muscle Volume (L)** | 8.55 ± 2.25[2,3] | 8.38 ± 2.21[1,3] | 7.77 ± 1.98[1,2] |
| **Thigh Muscle Volume Index (L/m²)** | 2.92 ± 0.54[3] | 2.92 ± 0.54[3] | 2.80 ± 0.51[1,2] |
| **Mid-thigh Muscle Volume (L)** | 2.30 ± 0.56[2,3] | 2.28 ± 0.55[1,3] | 2.14 ± 0.51[1,2] |
| **Iliopsoas Muscle Volume (L)** | 0.65 ± 0.17[2,3] | 0.63 ± 0.17[1,3] | 0.58 ± 0.15[12] |
| **Iliopsoas Muscle Volume Index (L/m²)** | 0.22 ± 0.04[3] | 0.22 ± 0.04[3] | 0.21 ± 0.04[1,2] |
| **Muscle quality IDPs** | | | |
| **Thigh IMAT Volume (L)** | 0.73 ± 0.31[2,3] | 0.82 ± 0.36[1,3] | 1.02 ± 0.46[1,2] |
| **Thigh IMAT Volume Index (mL/m²)** | 252.13 ± 100.63[2,3] | 288.46 ± 121.96[1,3] | 368.42 ± 152.03[1,2] |
| **Mid-thigh IMAT Volume (mL)** | 45.93 ± 39.49[2,3] | 57.50 ± 47.16[1,3] | 83.56 ± 62.98[1,2] |
| **Mid-thigh IMAT/Muscle (%)** | 1.98 ± 1.59[2,3] | 2.49 ± 1.93[1,3] | 3.73 ± 2.86[12] |
| **Paraspinal Muscle PDFF (%)** | 6.84 ± 3.23[2,3] | 7.96 ± 4.34[1,3] | 10.45 ± 6.00[1,2] |
| **Paraspinal Muscle Iron Concentration (mg/g)** | 1.20 ± 0.12 | 1.20 ± 0.12 | 1.18 ± 0.10 |

Values are reported as mean and standard deviation. One-way ANOVA was used to compare the following groups: not frail - pre frail, not frail - frail and pre frail - frail participants. The numbers 1, 2, and 3 indicate those variables that were statistically significantly different between groups after Bonferroni correction ($p = 8.6 \times 10^{-5}$). IMAT: Intermuscular adipose tissue; PDFF: Proton density fat fraction.

**Supplementary Table S8.** Regression coefficients between dynapenia and the anthropometric covariate and muscle and fat IDPs.

| | **Models - Dynapenia** | | | | | | |
|---|---|---|---|---|---|---|---|
| | **(1)** | **(2)** | **(3)** | **(4)** | **(5)** | **(6)** | **(7)** |
| **Age** | 1.527** | 1.673** | 1.551** | 1.672** | 1.558** | 1.582** | 1.715** |
| | (1.463, 1.594) | (1.604, 1.745) | (1.487, 1.619) | (1.603, 1.744) | (1.492, 1.628) | (1.515, 1.652) | (1.644, 1.789) |
| **Male** | 4.269** | 1.779** | 3.740** | 1.801** | 2.514** | 2.608** | 1.341** |
| | (3.688, 4.941) | (1.576, 2.008) | (3.262, 4.288) | (1.605, 2.020) | (2.220, 2.847) | (2.291, 2.969) | (1.205, 1.493) |
| **Asian** | 1.377* | 1.855** | 1.496* | 1.907** | 1.702* | 1.504* | 1.938** |
| | (1.026, 1.822) | (1.387, 2.445) | (1.115, 1.976) | (1.426, 2.512) | (1.271, 2.245) | (1.122, 1.986) | (1.449, 2.552) |
| **Black** | 0.607 | 0.567 | 0.680 | 0.633 | 0.682 | 0.453* | 0.456* |
| | (0.272, 1.163) | (0.255, 1.082) | (0.305, 1.305) | (0.284, 1.212) | (0.306, 1.305) | (0.204, 0.865) | (0.205, 0.870) |
| **Chinese** | 1.075 | 1.440 | 1.139 | 1.464 | 1.339 | 1.154 | 1.467 |
| | (0.554, 1.902) | (0.744, 2.544) | (0.587, 2.016) | (0.756, 2.588) | (0.691, 2.366) | (0.595, 2.043) | (0.757, 2.591) |
| **Others** | 0.892 | 1.016 | 0.935 | 1.043 | 1.020 | 0.875 | 0.980 |
| | (0.590, 1.299) | (0.674, 1.473) | (0.619, 1.359) | (0.692, 1.512) | (0.676, 1.481) | (0.580, 1.271) | (0.651, 1.419) |
| **Alcohol frequency [Daily]** | 0.681** | 0.662** | 0.693** | 0.672** | 0.684** | 0.673** | 0.664** |
| | (0.601, 0.770) | (0.585, 0.749) | (0.612, 0.784) | (0.594, 0.760) | (0.604, 0.774) | (0.594, 0.761) | (0.587, 0.750) |
| **Alcohol frequency [1-4 times per week]** | 0.765** | 0.746** | 0.774** | 0.754** | 0.766** | 0.764** | 0.748** |
| | (0.694, 0.844) | (0.677, 0.822) | (0.703, 0.854) | (0.685, 0.831) | (0.696, 0.845) | (0.694, 0.842) | (0.679, 0.824) |
| **Alcohol frequency [1-3 times per month]** | 0.780* | 0.772* | 0.785* | 0.777* | 0.784* | 0.784* | 0.775* |
| | (0.679, 0.895) | (0.672, 0.885) | (0.683, 0.901) | (0.677, 0.891) | (0.683, 0.899) | (0.683, 0.900) | (0.675, 0.888) |
| **Smoking status [Previous]** | 0.966 | 0.955 | 0.950 | 0.947 | 0.955 | 0.933 | 0.935 |
| | (0.891, 1.046) | (0.882, 1.034) | (0.877, 1.029) | (0.874, 1.025) | (0.881, 1.034) | (0.861, 1.010) | (0.863, 1.012) |
| | 0.996 | 1.001 | 0.949 | 0.967 | 0.958 | 0.960 | 0.984 |

| | | | | | | | |
|---|---|---|---|---|---|---|---|
| **Smoking status [Current]** | (0.794, 1.236) | (0.799, 1.240) | (0.756, 1.177) | (0.771, 1.198) | (0.764, 1.189) | (0.765, 1.190) | (0.785, 1.218) |
| **Townsend deprivation index** | 1.055* | 1.066* | 1.054* | 1.066* | 1.061* | 1.054* | 1.064* |
| | (1.016, 1.094) | (1.028, 1.106) | (1.016, 1.094) | (1.027, 1.106) | (1.023, 1.101) | (1.016, 1.094) | (1.025, 1.104) |
| **MET** | 0.846** | 0.838** | 0.849** | 0.843** | 0.842** | 0.836** | 0.830** |
| | (0.813, 0.880) | (0.806, 0.872) | (0.816, 0.884) | (0.810, 0.877) | (0.810, 0.876) | (0.804, 0.870) | (0.798, 0.863) |
| **Total Muscle Volume** | 0.392** | | | | | | |
| | (0.361, 0.426) | | | | | | |
| **Total Muscle Volume Index** | | 0.665** | | | | | |
| | | (0.623, 0.710) | | | | | |
| **Thigh Muscle Volume** | | | 0.416** | | | | |
| | | | (0.385, 0.449) | | | | |
| **Thigh Muscle Volume Index** | | | | 0.650** | | | |
| | | | | (0.611, 0.692) | | | |
| **Mid-thigh Muscle Volume** | | | | | 0.522** | | |
| | | | | | (0.487, 0.560) | | |
| **Iliopsoas Muscle Volume** | | | | | | 0.519** | |
| | | | | | | (0.482, 0.558) | |
| **Iliopsoas Muscle Volume Index** | | | | | | | 0.789** |
| | | | | | | | (0.745, 0.835) |
| **Constant** | 0.041** | 0.068** | 0.044** | 0.067** | 0.055** | 0.055** | 0.080** |
| | (0.037, 0.046) | (0.061, 0.076) | (0.039, 0.049) | (0.060, 0.074) | (0.049, 0.061) | (0.049, 0.061) | (0.072, 0.088) |
| **Observations** | 41,943 | 41,943 | 41,943 | 41,943 | 41,943 | 41,943 | 41,943 |
| **AIC** | 20854.5 | 21229.4 | 20857.8 | 21195.1 | 21038.9 | 21057.6 | 21318.7 |

| | **Models - Dynapenia** (Continued) | | | | | |
|---|---|---|---|---|---|---|
| | **(8)** | **(9)** | **(10)** | **(11)** | **(12)** | **(13)** |
| **Age** | 1.784** | 1.761** | 1.781** | 1.752** | 1.753** | 1.804** |
| | (1.713, 1.859) | (1.690, 1.835) | (1.710, 1.856) | (1.682, 1.826) | (1.681, 1.828) | (1.732, 1.880) |
| **Male** | 0.948 | 0.965 | 0.954 | 1.016 | 0.998 | 0.973 |
| | (0.878, 1.024) | (0.895, 1.041) | (0.885, 1.030) | (0.942, 1.097) | (0.925, 1.077) | (0.902, 1.049) |
| **Asian** | 2.197** | 2.157** | 2.191** | 2.127** | 2.149** | 2.179** |
| | (1.646, 2.887) | (1.616, 2.835) | (1.641, 2.879) | (1.592, 2.797) | (1.609, 2.825) | (1.633, 2.864) |
| **Black** | 0.437* | 0.427* | 0.428* | 0.430* | 0.449* | 0.452* |
| | (0.197, 0.833) | (0.192, 0.813) | (0.193, 0.816) | (0.193, 0.820) | (0.202, 0.854) | (0.203, 0.860) |
| **Chinese** | 1.657 | 1.706 | 1.664 | 1.690 | 1.646 | 1.590 |
| | (0.856, 2.926) | (0.881, 3.012) | (0.860, 2.937) | (0.873, 2.984) | (0.850, 2.908) | (0.821, 2.805) |
| **Others** | 1.015 | 1.013 | 1.018 | 1.023 | 1.010 | 1.002 |
| | (0.675, 1.470) | (0.673, 1.467) | (0.676, 1.473) | (0.679, 1.481) | (0.671, 1.463) | (0.666, 1.450) |
| **Alcohol frequency [Daily]** | 0.668** | 0.668** | 0.671** | 0.670** | 0.675** | 0.674** |
| | (0.591, 0.756) | (0.590, 0.755) | (0.593, 0.758) | (0.592, 0.758) | (0.597, 0.763) | (0.595, 0.762) |
| **Alcohol frequency [1-4 times per week]** | 0.746** | 0.749** | 0.748** | 0.752** | 0.751** | 0.746** |
| | (0.677, 0.822) | (0.680, 0.826) | (0.679, 0.824) | (0.682, 0.828) | (0.682, 0.827) | (0.678, 0.822) |
| **Alcohol frequency [1-3 times per month]** | 0.770* | 0.772* | 0.772* | 0.774* | 0.774* | 0.770* |
| | (0.671, 0.883) | (0.673, 0.885) | (0.672, 0.885) | (0.674, 0.888) | (0.674, 0.887) | (0.671, 0.882) |
| **Smoking status [Previous]** | 0.925 | 0.912* | 0.920* | 0.912* | 0.926 | 0.939 |
| | (0.854, 1.002) | (0.842, 0.988) | (0.849, 0.996) | (0.842, 0.988) | (0.855, 1.003) | (0.867, 1.016) |
| **Smoking status [Current]** | 0.997 | 0.984 | 0.997 | 0.986 | 0.998 | 1.004 |
| | (0.796, 1.234) | (0.786, 1.219) | (0.796, 1.234) | (0.787, 1.221) | (0.797, 1.236) | (0.802, 1.243) |

| | | | | | | |
|---|---|---|---|---|---|---|
| **Townsend deprivation index** | 1.063* | 1.058* | 1.060* | 1.057* | 1.061* | 1.066* |
| | (1.024, 1.103) | (1.020, 1.098) | (1.022, 1.100) | (1.019, 1.097) | (1.023, 1.101) | (1.027, 1.106) |
| **MET** | 0.831** | 0.843** | 0.836** | 0.844** | 0.833** | 0.821** |
| | (0.799, 0.865) | (0.810, 0.877) | (0.803, 0.869) | (0.811, 0.879) | (0.801, 0.866) | (0.790, 0.854) |
| **Thigh IMAT Volume** | 1.060* | | | | | |
| | (1.022, 1.098) | | | | | |
| **Thigh IMAT Volume Index** | | 1.129** | | | | |
| | | (1.091, 1.168) | | | | |
| **Mid-thigh IMAT Volume** | | | 1.099** | | | |
| | | | (1.062, 1.136) | | | |
| **Mid-thigh IMAT/Muscle** | | | | 1.162** | | |
| | | | | (1.123, 1.201) | | |
| **Paraspinal Muscle PDFF** | | | | | 1.094** | |
| | | | | | (1.059, 1.130) | |
| **Paraspinal Muscle Iron Concentration** | | | | | | 1.057* |
| | | | | | | (1.021, 1.094) |
| **Constant** | 0.096** | 0.095** | 0.095** | 0.092** | 0.093** | 0.094** |
| | (0.087, 0.104) | (0.086, 0.103) | (0.087, 0.104) | (0.084, 0.100) | (0.085, 0.101) | (0.086, 0.103) |
| **Observations** | 41,943 | 41,943 | 41,943 | 41,943 | 41,943 | 41,943 |
| **AIC** | 21376.3 | 21339.9 | 21357.6 | 21313.7 | 21357.1 | 21376.3 |

Values are presented as odds ratios (OR) and 95% confidence intervals (CI) in parentheses. * indicate statistically significant for $p < 0.05$, ** indicate statistically significant after Bonferroni correction ($p = 8.6 \times 10^{-5}$). Models 1-7 were adjusted for age, sex, ethnicity, alcohol intake frequency, smoking status, Townsend deprivation index, MET, as well as each muscle IDPs, separately. *(1) - Total Muscle Volume; (2) - Total Muscle Volume Index; (3) - Thigh Muscle Volume; (4) - Thigh Muscle Volume Index; (5) - Iliopsoas Muscle Volume; (6) - Mid-thigh Muscle Volume; (7) - Iliopsoas Muscle Volume Index; (8) - Thigh IMAT Volume; (9) - Thigh IMAT Volume Index; (10) - Mid-thigh IMAT Volume; (11) - Mid-thigh IMAT/Muscle; (12) - Paraspinal Muscle PDFF; (13) - Paraspinal Muscle Iron Concentration.

**Supplementary Table S9.** Regression coefficients between frailty and the anthropometric covariate and muscle and fat IDPs.

| | Models - Frailty | | | | | | |
|---|---|---|---|---|---|---|---|
| | **(1)** | **(2)** | **(3)** | **(4)** | **(5)** | **(6)** | **(7)** |
| **Age** | 1.179** (1.157, 1.201) | 1.234** (1.212, 1.256) | 1.167** (1.145, 1.189) | 1.215** (1.193, 1.236) | 1.199** (1.176, 1.222) | 1.159** (1.137, 1.181) | 1.206** (1.185, 1.228) |
| **Male** | 1.228** (1.148, 1.309) | 0.841** (0.774, 0.908) | 1.336** (1.261, 1.412) | 0.960 (0.896, 1.024) | 1.058 (0.988, 1.127) | 1.373** (1.300, 1.446) | 1.015 (0.955, 1.076) |
| **Asian** | 1.473* (1.277, 1.669) | 1.646** (1.451, 1.842) | 1.445* (1.249, 1.640) | 1.582** (1.387, 1.777) | 1.547** (1.351, 1.742) | 1.398* (1.202, 1.595) | 1.567** (1.371, 1.762) |
| **Black** | 1.295* (1.038, 1.552) | 1.116 (0.859, 1.373) | 1.367* (1.109, 1.625) | 1.172 (0.914, 1.429) | 1.246 (0.989, 1.503) | 1.262 (1.005, 1.518) | 1.228 (0.973, 1.484) |
| **Chinese** | 1.001 (0.632, 1.369) | 1.094 (0.726, 1.462) | 0.980 (0.611, 1.349) | 1.065 (0.697, 1.433) | 1.047 (0.679, 1.415) | 0.957 (0.588, 1.326) | 1.057 (0.689, 1.425) |
| **Others** | 1.160 (0.956, 1.364) | 1.155 (0.952, 1.359) | 1.164 (0.960, 1.368) | 1.160 (0.956, 1.364) | 1.170 (0.966, 1.373) | 1.130 (0.926, 1.334) | 1.170 (0.966, 1.374) |
| **Alcohol frequency [Daily]** | 0.541** (0.471, 0.612) | 0.543** (0.473, 0.614) | 0.543** (0.472, 0.614) | 0.541** (0.470, 0.611) | 0.541** (0.471, 0.612) | 0.540** (0.470, 0.611) | 0.541** (0.471, 0.612) |
| **Alcohol frequency [1-4 times per week]** | 0.618** (0.562, 0.675) | 0.617** (0.560, 0.674) | 0.621** (0.564, 0.677) | 0.616** (0.560, 0.673) | 0.618** (0.561, 0.674) | 0.620** (0.564, 0.677) | 0.617** (0.560, 0.674) |
| **Alcohol frequency [1-3 times per month]** | 0.794** (0.718, 0.870) | 0.790** (0.714, 0.866) | 0.796** (0.719, 0.872) | 0.791** (0.715, 0.867) | 0.792** (0.716, 0.869) | 0.796** (0.720, 0.872) | 0.792** (0.716, 0.868) |
| **Smoking status [Previous]** | 1.156** (1.112, 1.200) | 1.142** (1.099, 1.186) | 1.156** (1.113, 1.200) | 1.149** (1.105, 1.192) | 1.152** (1.108, 1.195) | 1.152** (1.109, 1.196) | 1.151** (1.107, 1.194) |
| | 1.277** | 1.273** | 1.268** | 1.279** | 1.274** | 1.268** | 1.276** |

| | | | | | | | |
|---|---|---|---|---|---|---|---|
| **Smoking status [Current]** | (1.163, 1.391) | (1.159, 1.387) | (1.154, 1.382) | (1.165, 1.392) | (1.161, 1.388) | (1.154, 1.382) | (1.162, 1.390) |
| **Townsend deprivation index** | 1.056** | 1.057** | 1.055** | 1.057** | 1.057** | 1.054** | 1.057** |
| | (1.035, 1.077) | (1.036, 1.078) | (1.035, 1.076) | (1.036, 1.078) | (1.036, 1.077) | (1.034, 1.075) | (1.036, 1.078) |
| **MET** | 0.865** | 0.855** | 0.867** | 0.859** | 0.862** | 0.865** | 0.861** |
| | (0.844, 0.886) | (0.834, 0.876) | (0.846, 0.888) | (0.837, 0.880) | (0.841, 0.883) | (0.844, 0.886) | (0.840, 0.882) |
| **Total Muscle Volume** | 0.905** | | | | | | |
| | (0.864, 0.945) | | | | | | |
| **Total Muscle Volume Index** | | 1.141** | | | | | |
| | | (1.107, 1.174) | | | | | |
| **Thigh Muscle Volume** | | | 0.858** | | | | |
| | | | (0.820, 0.896) | | | | |
| **Thigh Muscle Volume Index** | | | | 1.051* | | | |
| | | | | (1.019, 1.083) | | | |
| **Mid-thigh Muscle Volume** | | | | | 0.987 | | |
| | | | | | (0.951, 1.022) | | |
| **Iliopsoas Muscle Volume** | | | | | | 0.842** | |
| | | | | | | (0.805, 0.879) | |
| **Iliopsoas Muscle Volume Index** | | | | | | | 1.013 |
| | | | | | | | (0.983, 1.044) |
| **Not Frail\|Pre Frail** | 1.241** | 1.028 | 1.296** | 1.096* | 1.152** | 1.309** | 1.129** |
| | (1.238, 1.243) | (1.028, 1.028) | (1.293, 1.299) | (1.095, 1.097) | (1.15, 1.153) | (1.305, 1.312) | (1.127, 1.13) |
| **Pre Frail\|Frail** | 75.47** | 62.636** | 78.958** | 66.648** | 69.984** | 79.821** | 68.579** |
| | (72.37, 78.702) | (60.173, 65.201) | (75.682, 82.375) | (63.988, 69.418) | (67.159, 72.928) | (76.502, 83.285) | (65.824, 71.45) |

| | | | | | | | |
|---|---|---|---|---|---|---|---|
| **Observations** | 40,841 | 40,841 | 40,841 | 40,841 | 40,841 | 40,841 | 40,841 |
| **AIC** | 57565.3 | 57529.9 | 57526.8 | 57579.7 | 57588.3 | 57505.4 | 57588.1 |

| | **Models - Frailty** (Continued) | | | | | |
|---|---|---|---|---|---|---|
| | **(8)** | **(9)** | **(10)** | **(11)** | **(12)** | **(13)** |
| **Age** | 1.163** | 1.141** | 1.172** | 1.143** | 1.124** | 1.203** |
| | (1.142, 1.184) | (1.120, 1.163) | (1.150, 1.193) | (1.122, 1.164) | (1.103, 1.146) | (1.182, 1.223) |
| **Male** | 0.929* | 1.027 | 0.993 | 1.148** | 1.128** | 1.035 |
| | (0.887, 0.971) | (0.985, 1.068) | (0.951, 1.034) | (1.106, 1.190) | (1.086, 1.170) | (0.994, 1.076) |
| **Asian** | 1.552** | 1.477* | 1.535** | 1.454* | 1.467* | 1.557** |
| | (1.356, 1.748) | (1.280, 1.673) | (1.339, 1.731) | (1.257, 1.650) | (1.270, 1.664) | (1.362, 1.752) |
| **Black** | 1.154 | 1.127 | 1.142 | 1.203 | 1.275 | 1.231 |
| | (0.896, 1.413) | (0.868, 1.386) | (0.882, 1.401) | (0.944, 1.461) | (1.017, 1.533) | (0.975, 1.486) |
| **Chinese** | 1.245 | 1.236 | 1.178 | 1.170 | 1.160 | 1.052 |
| | (0.876, 1.614) | (0.867, 1.606) | (0.809, 1.547) | (0.801, 1.540) | (0.790, 1.530) | (0.684, 1.420) |
| **Others** | 1.197 | 1.177 | 1.185 | 1.194 | 1.192 | 1.169 |
| | (0.992, 1.402) | (0.971, 1.383) | (0.980, 1.390) | (0.989, 1.399) | (0.987, 1.396) | (0.965, 1.373) |
| **Alcohol frequency [Daily]** | 0.522** | 0.522** | 0.530** | 0.529** | 0.542** | 0.541** |
| | (0.451, 0.593) | (0.450, 0.593) | (0.459, 0.601) | (0.458, 0.601) | (0.470, 0.613) | (0.470, 0.611) |
| **Alcohol frequency [1-4 times per week]** | 0.615** | 0.617** | 0.618** | 0.620** | 0.625** | 0.617** |
| | (0.558, 0.672) | (0.559, 0.674) | (0.561, 0.675) | (0.563, 0.678) | (0.568, 0.682) | (0.560, 0.674) |
| **Alcohol frequency [1-3 times per month]** | 0.789** | 0.790** | 0.791** | 0.795** | 0.798** | 0.792** |
| | (0.712, 0.866) | (0.714, 0.867) | (0.715, 0.868) | (0.718, 0.871) | (0.721, 0.874) | (0.716, 0.868) |
| **Smoking status [Previous]** | 1.094** | 1.085* | 1.102** | 1.101** | 1.118** | 1.151** |
| | (1.050, | (1.041, | (1.058, | (1.057, | (1.074, 1.162) | (1.107, 1.194) |

| | | | | | | |
|---|---|---|---|---|---|---|
| | 1.138) | 1.129) | 1.146) | 1.145) | | |
| **Smoking status [Current]** | 1.211* | 1.202* | 1.230* | 1.222* | 1.249* | 1.276** |
| | (1.096, 1.325) | (1.087, 1.317) | (1.116, 1.345) | (1.108, 1.337) | (1.134, 1.363) | (1.162, 1.389) |
| **Townsend deprivation index** | 1.047** | 1.042* | 1.045** | 1.043** | 1.045** | 1.057** |
| | (1.026, 1.067) | (1.021, 1.063) | (1.024, 1.065) | (1.022, 1.064) | (1.024, 1.066) | (1.036, 1.078) |
| **MET** | 0.900** | 0.908** | 0.895** | 0.902** | 0.888** | 0.861** |
| | (0.879, 0.922) | (0.887, 0.929) | (0.874, 0.917) | (0.881, 0.923) | (0.867, 0.910) | (0.841, 0.882) |
| **Thigh IMAT Volume** | 1.328** | | | | | |
| | (1.306, 1.350) | | | | | |
| **Thigh IMAT Volume Index** | | 1.396** | | | | |
| | | (1.374, 1.418) | | | | |
| **Mid-thigh IMAT Volume** | | | 1.349** | | | |
| | | | (1.327, 1.372) | | | |
| **Mid-thigh IMAT/Muscle** | | | | 1.408** | | |
| | | | | (1.383, 1.432) | | |
| **Paraspinal Muscle PDFF** | | | | | 1.350** | |
| | | | | | (1.327, 1.372) | |
| **Paraspinal Muscle Iron Concentration** | | | | | | 0.995 |
| | | | | | | (0.975, 1.015) |
| **Not Frail\|Pre Frail** | 1.054 | 1.104* | 1.095* | 1.175** | 1.183** | 1.139** |
| | (1.053, 1.054) | (1.102, 1.105) | (1.094, 1.096) | (1.173, 1.177) | (1.181, 1.185) | (1.138, 1.14) |
| **Pre Frail\|Frail** | 66.512** | 70.934** | 69.906** | 75.93** | 75.815** | 69.211** |
| | (63.859, 69.276) | (68.062, 73.927) | (67.085, 72.845) | (72.808, 79.186) | (72.699, 79.066) | (66.425, 72.114) |
| **Observations** | 40,841 | 40,841 | 40,841 | 40,841 | 40,841 | 40,841 |
| **AIC** | 56936.6 | 56672.4 | 56892.5 | 56787.1 | 56861.3 | 57588.6 |

Values are presented as odds ratios (OR) and 95% confidence intervals (CI) in parentheses. * indicate statistically significant for $p < 0.05$, ** indicate statistically significant after Bonferroni correction ($p = 8.6 \times 10^{-5}$). Models 1-7 were adjusted for age, sex, ethnicity, alcohol intake frequency, smoking status, Townsend deprivation index, MET, as well as each muscle IDPs, separately. *(1) - Total Muscle Volume; (2) - Total Muscle Volume Index; (3) - Thigh Muscle Volume; (4) - Thigh Muscle Volume Index; (5) - Iliopsoas Muscle Volume; (6) - Mid-thigh Muscle Volume; (7) - Iliopsoas Muscle Volume Index; (8) - Thigh IMAT Volume; (9) - Thigh IMAT Volume Index; (10) - Mid-thigh IMAT Volume; (11) - Mid-thigh IMAT/Muscle; (12) - Paraspinal Muscle PDFF; (13) - Paraspinal Muscle Iron Concentration.

**Supplementary Table S10.** Performance of the logistic regression models for the associations between dynapenia or frailty and muscle volume and quality IDPs adjusted for the anthropometric covariates.

| | **Dynapenia**<br>AUC (95%CI) | **Frailty**<br>C-index |
|---|---|---|
| **Muscle volume IDPs** | | |
| **Total Muscle Volume (L)** | **0.707 (0.698, 0.716)** | 0.590 |
| **Total Muscle Volume Index (L/m²)** | 0.682 (0.673, 0.691) | 0.593 |
| **Thigh Muscle Volume (L)** | 0.707 (0.697, 0.716) | 0.593 |
| **Thigh Muscle Volume Index (L/m²)** | 0.684 (0.675, 0.694) | 0.590 |
| **Mid-thigh Muscle Volume (L)** | 0.695 (0.685, 0.704) | 0.590 |
| **Iliopsoas Muscle Volume (L)** | 0.695 (0.686, 0.704) | 0.594 |
| **Iliopsoas Muscle Volume Index (L/m²)** | 0.676 (0.666, 0.685) | 0.590 |
| **Muscle quality IDPs** | | |
| **Thigh IMAT Volume (L)** | 0.672 (0.662, 0.681) | 0.613 |
| **Thigh IMAT Volume Index (mL/m²)** | 0.674 (0.665, 0.684) | **0.621** |
| **Mid-thigh IMAT Volume (mL)** | 0.673 (0.664, 0.683) | 0.616 |
| **Mid-thigh IMAT/Muscle (%)** | 0.677 (0.667, 0.686) | 0.618 |
| **Paraspinal Muscle PDFF (%)** | 0.674 (0.664, 0.683) | 0.614 |
| **Paraspinal Muscle Iron Concentration (mg/g)** | 0.672 (0.662, 0.681) | 0.590 |

The performance of dynapenia is presented by the area under the curve (AUC) with 95% confidence intervals (CIs) in parentheses, while the performance of frailty is measured by the concordance index (c-index). Values with the highest AUC and c-index for each health condition are shown in bold. IMAT: Intermuscular adipose tissue; PDFF: Proton density fat fraction.

### 1 References

Fried, L.P. *et al.* (2001) ‘Frailty in older adults: evidence for a phenotype’, *The journals of gerontology. Series A, Biological sciences and medical sciences*, 56(3), pp. M146–56.
